# Assessment of the Impacts of Pharmaceutical and Non-pharmaceutical Intervention on COVID-19 in South Africa Using Mathematical Model

**DOI:** 10.1101/2020.11.13.20231159

**Authors:** Rabiu Musa, Absalom E. Ezugwu, Godwin C. E. Mbah

## Abstract

The novel coronal virus has spread across more than 213 countries within the space of six months causing devastating public health hazard and monumental economic loss. In the absence of clinically approved pharmaceutical intervention, attentions are shifted to non-pharmaceutical controls to mitigate the burden of the novel pandemic. In this regard, a ten mutually exclusive compartmental mathematical model is developed to investigate possible effects of both pharmaceutical and non-pharmaceutical controls incorporating both private and government’s quarantine and treatments. Several reproduction numbers were calculated and used to determine the impact of both control measures as well as projected benefits of social distancing, treatments and vaccination. We investigate and compare the possible impact of social distancing incorporating different levels of vaccination, with vaccination programme incorporating different levels of treatment. Using the officially published South African COVID-19 data, the numerical simulation shows that the community reproduction threshold will be 30 when there is no social distancing but will drastically reduced to 5 (about 83% reduction) when social distancing is enforced. Furthermore, when there is vaccination with perfect efficacy, the community reproduction threshold will be 4 which increases to 12 (about 67% increment) without vaccination. We also established that the implementation of both interventions is enough to curtail the spread of COVID-19 pandemic in South Africa which is in confirmation with the recommendation of the world health organization.

## 1 Introduction

The novel corona-virus which was first discovered in Wuhan city of China in December 2019 has taken the world by storm [27, 29]. Presently, COVID-19 (caused by the novel SARS-CoV-2 corona-virus) has affected over 213 countries across the world with more than 18 million cases and daily reported cases of 250 000 as at August 5, 2020 [4, 27, 29]. The United State maintains its lead as the epicenter of the pandemic with more than 4,000,000 cases and about 160,0000 causalities. In Africa, South Africa is the epicenter of the pandemic with over half a million cases, more than 8,000 deaths but with an impressive 69% recovery rate as at August 4, 2020 [7].

The National Institute for Communicable Diseases confirmed the first case of COVID-19 in South Africa on March 1, 2020 [19]. Majority of the COVID-19 fatality and severe cases occur among the aged populations of about 60 years and above, and to people with underlying medical conditions. However, adults and young people with sound medical conditions are less prone to COVID-19 fatality [2].

Just like other viruses such as the Flu and Severe Acute respiratory Syndrome diseases [29, 33], COVID-19 is also transmitted from person-to-person through direct contact with infected surfaces, objects, body fluid, blood or through inhalation of respiratory droplets from infected individuals whether symptomatic or asymptomatic [1]. The latent period of the virus is between 2-14 days [17, 5]. Majority of the infected persons show mild symptom (or no symptoms at all) though popular symptoms include but not limited to headache, fever, shortness of breath, cough and pneumonia ranging from mild to severe cases [27].

Due to the unavailability of pharmaceutical interventions such as suitable vaccines and a COVID-19 anti-viral medication, the implementation of non-pharmaceutical interventions like restriction of movement, social distancing, face-masks usage, quarantine of exposed persons, isolation of confirmed and tested cases, contact-tracing and so on are the commonly prescribed measures by the world health organization [8, 20]. The recommendation of strict social distancing measure was also among the regular healthcare guidelines that was prescribed by the South African medical practitioners for curbing the spread of COVID-19, since the virus is transmitted among persons. For this reason, approximately five months ago, strict social distancing intervention like complete lock-down, school closure, closure of non-essential businesses were all imposed in several countries of the world including South Africa which was later relaxed in mid-May, 2020 [24, 28].

Several researchers, government agencies and pharmaceutical companies have stormed the laboratories to develop safe and effective vaccines to curtail the menace of COVID-19. Realistically, development of a vaccine generally takes more than a year, many organizations have already started and all hopes are high that the vaccine will be available sooner rather than later [3]. Fortunately at the Oxford University, the Jenner Institute among other research institutes have developed a new COVID-19 vaccine which is expected to be available before the end of 2020 [14].

Several models have been published on COVID-19 [8, 10, 11, 17, 18, 32], some of them are reviewed subsequently. Gumel et al. (2020) [11] studied the possibility of an imperfect COVID-19 vaccine in the United State of America. In their work, the equation for the minimum number of susceptible individuals (that are excluded from the vaccination programme) needed to be vaccinated to obtain vaccine-induced community herd immunity was developed. The authors further investigated the epidemiological consequence of the herd immunity threshold and stressed that the virus can be totally controlled or defeated if the minimum herd immunity threshold is achieved in the country. They finally simulated the model equation using certain parameter values for the concerned states in the US, including, Florida and New York and determined the minimum herd immunity threshold for each state.

Jia et al. (2020) [12] studied the control of COVID-19 and the impact of policy interventions with meteorological factors in China using seven compartments. After fitting their model with officially published data, they estimated parameter values for the proposed model using Least-Squares procedure. After obtaining the reproduction number of each province in China, they proposed that the quarantine period must be long enough to reduce contact. They also introduced comprehensive meteorological index to represent the meteorological factors as well as vaccination strategies for different control measures.

Mushayabasa et al. (2020) [18] studied the role of governmental action and individual reaction on COVID-19 dynamics in South Africa using mathematical modeling approach. The proposed model examines relevant biological factors and effects of individual behavioral reaction including some interventions like consistent hygiene measures, quarantine, lock-down among others. Using officially published South African COVID-19 data available as at March 2020 to early May 2020, the model examines the optimal conditions necessary for the infection to die out and persist. Other commendable research works can be found in [15, 16, 34] and references therein.

The current study examines the effect of both pharmaceutical and non-pharmaceutical controls in curbing the infectiousness of the novel coronavirus disease. The incorporated pharmaceutical interventions include a perfect vaccine with varying potency and efficacy level for the susceptible individuals. Due to the over saturated nature of both the private and public hospitals because of the rapidly growing number of confirmed cases, the model considered personal treatment as an alternative to the South African government’s provided treatments. The same reason also applies to the already congested and over-subscribed isolation and quarantine centers provided by the South African government; we include self isolation mechanism as a non-pharmaceutical control techniques. Since no research has shown that some people are permanently immune from COVID-19, we considered that the infected individuals can become susceptible immediately after recovery. Furthermore, we considered the effect of social distancing and its efficacy level as well as vaccination. All these underline the novelty of this research work because to the best of our knowledge, no such model has considered both pharmaceutical and non-pharmaceutical control mechanism concurrently. To examine the reliability of the model, the model parameters were parameterized using officially published South African COVID-19 data that provide a realistic assessment and estimation of the severity of the virus in the country population.

The rest of the paper is organized as follows. Section 2 discussed materials and methods used in the analysis as well as the model formulation. The main analytic results are discussed in section 3. Section 4 entails the numerical simulation and the projected impacts of the interventions while section 5 discussed and compared results with the existing literature findings. Finally, the paper ends with a concluding remark in section 6.

## 2 Mathematical Model Formulation

The novel COVID-19 model we proposed consists of 10 mutually exclusive compartments of susceptible class *S*(*t*), vaccinated susceptible class *S*_*v*_(*t*), unvaccinated susceptible class *S*_*uv*_(*t*), Exposed class *E*(*t*), individuals on government’s quarantined facility *G*_*Q*_(*t*), individuals on personal quarantine facility *P*_*Q*_(*t*), infected individuals *I*(*t*), individuals on personal treatment (*P*_*T*_), individuals on government’s treatment facility *G*_*T*_ (*t*) and the recovered class *R*(*t*) such that the total population is given by

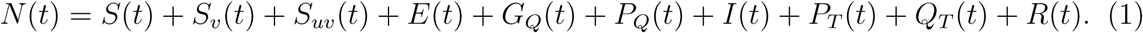

We need to declare here that Λ is the recruitment rate of individuals into the community. The susceptible class is reduced by a proportion *ρ* for the vaccinated susceptible individuals who move directly into the *S*_*v*_ class while a proportion (1 − *ρ*) who are not vaccinated perhaps due to lack of information or fear of contracting the virus move to the *S*_*uv*_(*t*) class. The proportion of people who are found uninfected after the 14-day governments’ quarantine move to the *S* class at the rate *b*_2_, *e* is the rate at which vaccinated susceptible persons loses their immunity and return to the susceptible class.

Those who are equally found uninfected after the 14-day personal quarantine also move to the susceptible class at the rate *b*_1_, the recovered persons after treatment or after being tested negative loss their immunity and hence return back to the susceptible class at the rate *η*_2_. The susceptible class is further reduced through natural death *μ* and breakthrough of infection after successfully contacting an infected individuals leading to infection at the rate *λ* given by

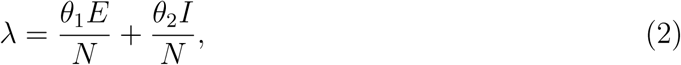

where *θ*_1_, *θ*_2_ is the contact rate for the exposed class *E* and infected class *I* respectively through contact with infected surface, droplet and infected individuals who failed to practice social distancing. It is worth noting here that we only considered infection can be contracted by individuals in classes *E* and *I* only because all other infected classes are either on treatment or quarantined and are assumed to be less involved in the transmission process. The unvaccinated susceptible class *S*_*uv*_ is reduced by breakthrough of infection denoted by *λ* and consequently moved to the *I* class.

The exposed class *E* is composed of susceptible individuals who are infected at the rate *λS* and vaccinated susceptible individuals at the rate *λS*_*v*_ who have been infected after successful contact with individuals who tested positive. They are reduced by movement of individuals at the rate *v*_1_ in to *G*_*Q*_ class, *v*_2_ into *I* class, *v*_3_ into *P*_*Q*_ class and natural death at the rate *μ*. We should also mention here that the exposed individuals also include those with mild symptoms of COVID-19, this can be confirmed and affirmed by available published data from WHO that around 80% of the COVID-19 confirmed and tested cases show mild or no symptoms [30]. More so, persons in this class especially those who are 65 years and above or individuals suffering from underlying ailment can develop a mild form of pneumonia that are expected to self-isolate for minimum period of 14 days [23, 32]. Furthermore, majority of persons in this class whether with symptoms or not can be diagnosed through effective testing and contact tracing of the contacts of confirmed COVID-19 cases which consequently move to *I*(*t*) class, quarantined or treated.

The individuals in government’s quarantine facility *G*_*Q*_ are further reduced by progression rate *h*_1_ to *I* class and natural death. Those on personal quarantine facility are further reduced by progression rate *h*_2_ into *I* class. Also the quarantined individuals are those who have been traced following effective contact with a close contact who tested positive (i.e., a confirmed COVID-19 infected person they have been exposed to). Those quarantined are considered susceptible or unaware infected and are expected to be tested to determine their status. More so, people can also be quarantined because of either traveling to areas with high risk of infection such as China, Italy, Spain etc.

The *I* class contains progression from *E* class at the rate *v*_2_, *G*_*Q*_ class at the rate *h*_1_, *P*_*T*_ class at the rate *h*_2_, force of infection from *S*_*uv*_ at the rate *λS*_*uv*_ and the class is reduced by treatment rates *f*_1_ and *f*_2_, death rate *μ* and COVID-19 induced death rate *δ*. The compartment *P*_*T*_ and *G*_*T*_ consist of progression rates *f*_1_ and *f*_2_ respectively from *I* class, COVID-19 induced death rates *δ*_2_ and *δ*_1_ respectively and recovery rates *τ*_2_ and *τ*_1_ respectively while *η*_2_ is the loss of immunity rate after recovery.

The model is given by the following autonomous nonlinear differential equations

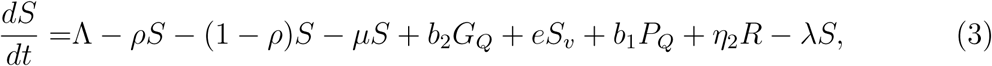

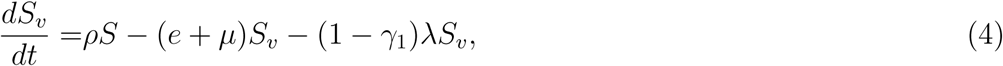

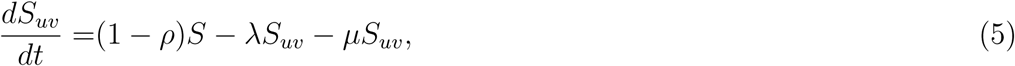

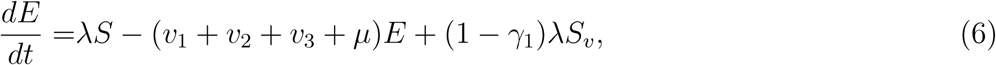

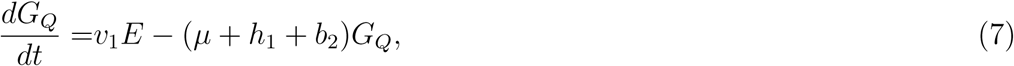

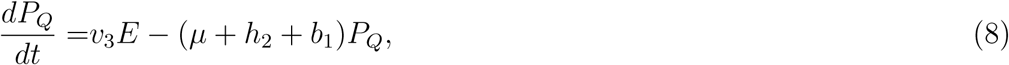

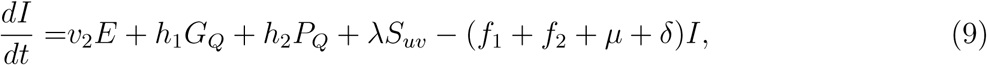

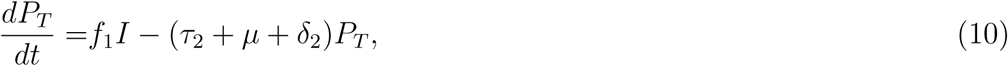

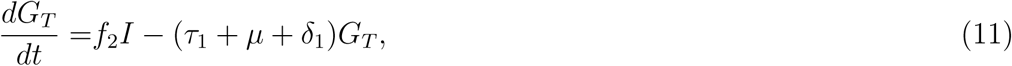

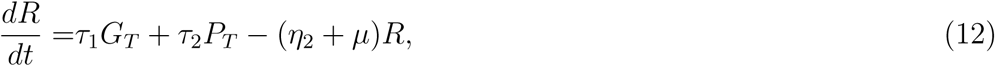

under the following initial conditions

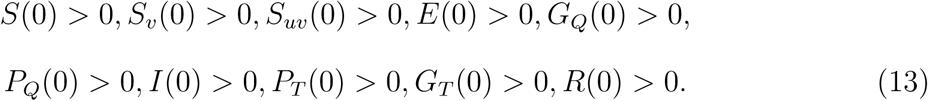

## 3 Analytic Results

### 3.1 Positivity of Solution

This section contains basic results of the model equations (3)-(12) established as follows.

#### Lemma 3.1.

*Let the initial conditions in* (13) *exist in the interval t* ∈ [0, ∞] *then the solutions S*(*t*), *S*_*v*_(*t*), *S*_*uv*_(*t*), *E*(*t*), *G*_*Q*_(*t*), *P*_*Q*_(*t*), *I*(*t*), *P*_*T*_ (*t*), *G*_*T*_ (*t*), *and R*(*t*) *of the model* *equations* (3)*-*(12) *are positive for all t* ≥ 0.

*Proof*. Vividly, it can be observed that the right-hand side of (3)-(12) is locally Liptschitzian, continuous and differentiable on the space of continuous function *κ*, the solution of *S*(*t*), *S*_*v*_(*t*), *S*_*uv*_(*t*), *E*(*t*), *G*_*Q*_(*t*), *P*_*Q*_(*t*), *I*(*t*), *P*_*T*_ (*t*), *G*_*T*_ (*t*), and *R*(*t*) under the stated initial conditions (13) uniquely exist on [0, *β*], where 0 < *β* ≤ ∞. Assuming that the solution components of the model equations (3)-(12) are not positive, then there exists a first time *t*^∗^ > 0 defined as follows

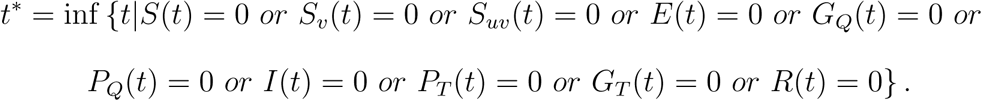

Suppose

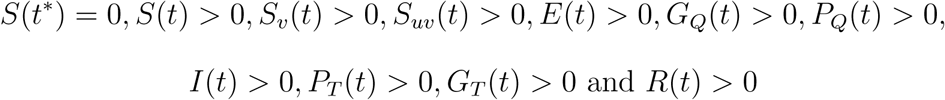

for *t* ∈ (0, *t*^∗^) then 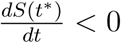. But unfortunately, from (3), 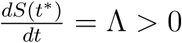 (where all model parameters are assumed positive) which shows that *S*(*t*) > 0. Consequently, following the same approach, we claim that the solutions *S*_*v*_(*t*), *S*_*uv*_(*t*), *E*(*t*), *G*_*Q*_(*t*), *P*_*Q*_(*t*), *I*(*t*), *P*_*T*_ (*t*), *G*_*T*_ (*t*) and *R*(*t*) are positive for all time *t* ≥ 0 [9, 21, 22].

### 3.2 Invariant Region

#### Lemma 3.2.

*The biological invariant region*

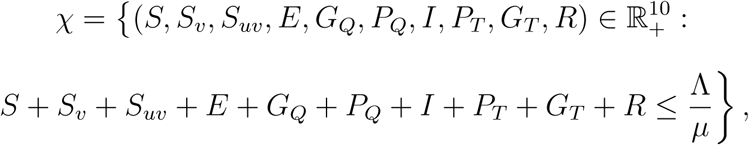

*is positively attracting and invariant for the model* (3)*-*(12).

*Proof*. The summation of equations (3)-(12) yields

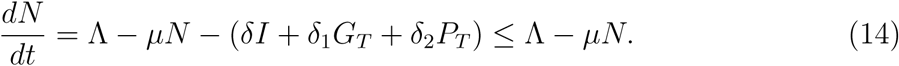

Using Gronwall inequality, the solution of (14) is given by

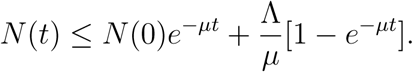

It can be observed that 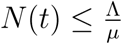 if 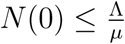. Therefore, *χ* is positively invariant. Hence, it is sufficient to study the COVID-19 model in *χ*. Thus, we conclude that the model (3)-(12) is epidemiologically welposed and attracting [9, 21, 22].

□

### 3.3 Local Stability of Virus-Free equilibrium (VFE)

The virus-free equilibrium of (3)-(12) is given by

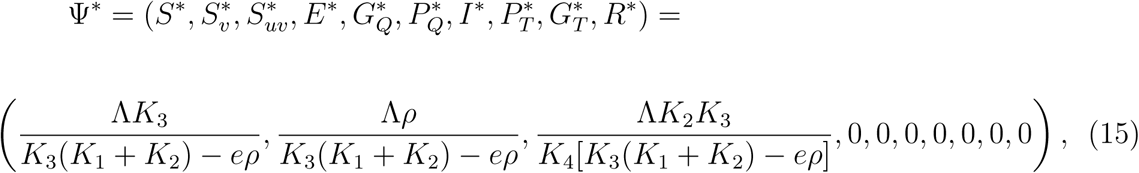

where 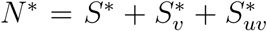. The linear stability of Ψ^∗^ can be carried out using the next generation matrix operator [6, 26] on (3)-(12). The new infection term ℱ and the transfer term 𝒱 are expressed as

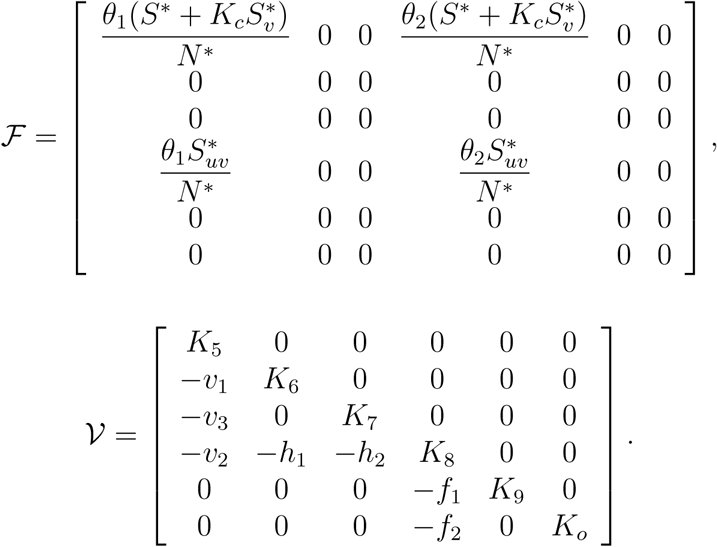

Using *ρ* as the spectral radius (magnitude of the dominate eigenvalue) of the next generation matrix ℱ 𝒱^−1^, the reproduction number is given by

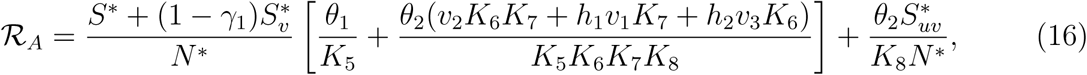

where

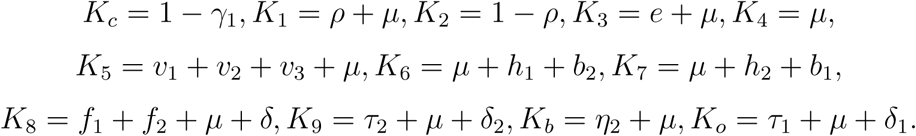

The threshold parameter ℛ_*A*_ is the control reproduction number of (3)-(12). It is used in measuring the average number of new COVID-19 cases generated by a single infected individual introduced into a community/population where a certain fraction of the population is under vaccination.

Furthermore, it can also be interpreted as the constituent reproduction number associated with the new COVID-19 cases generated by both exposed individuals *E* and infected individuals *I* expressed as

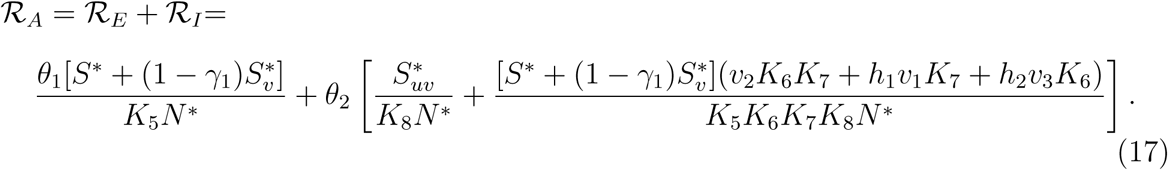

Hence, we have the result below.

#### Lemma 3.3.

*The VFE of the COVID-19 model* (3)*-*(12) *is locally asymptotically stable* (*LAS*) *if* ℛ_*A*_ < 1 *and unstable if* ℛ_*A*_ > 1.

The proof is standard and can be established using theorem 2 of [26].

### 3.4 Interpretation of the Reproduction Number

We re-express equation (17) as

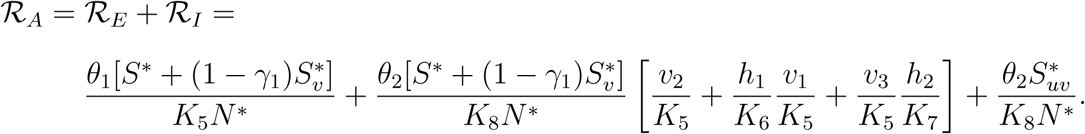

The constituent reproduction number ℛ_*E*_ can be interpreted as the product of the infection rate of susceptible and vaccinated susceptible individuals by the exposed humans near the VFE given by

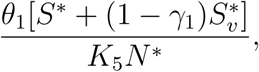

where the average period of time spent by the exposed humans in the exposed compartment *E* is given by 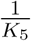.

The constituent reproduction number ℛ_*I*_ can be interpreted in component as follows. The proportion *v*_2_ of exposed individuals that survive the incubation period progresses to the infected class *I* after spending an average period of 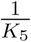 in *E* given by 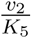 while that of *v*_1_ is the proportion of exposed individuals quarantined in a government facility after spending a period of 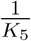 in *E* expressed as 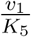. The remaining proportion *v*_3_ goes back home to self-quarantined after spending a period of 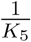 given by 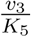.

The expression 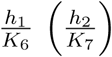 is for the proportion of individuals that progressed to the *I* class after 14-day period in government’s quarantine facility (personal quarantine facility) respectively. The infection rate of susceptible, vaccinated susceptible and unvaccinated susceptible individuals by the infected humans near the VFE is given by

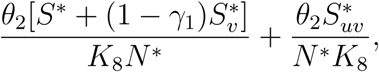

where, as mentioned earlier, the average period of time spent in the infected compartment *I* is given by 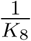. Thus, the summation ℛ_*E*_ + ℛ_*I*_ = ℛ_*A*_.

### 3.5 Impacts of the Control Measures on the Reproduction Number

Furthermore, in the absence of any pharmaceutical interventions ie. setting all pharmaceutical intervention parameters *ρ, γ*_1_, *f*_1_ and *f*_2_ to zero in (17), we have

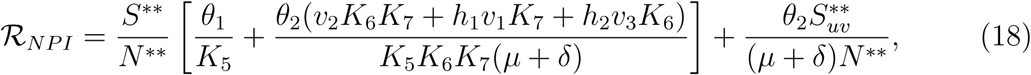

where

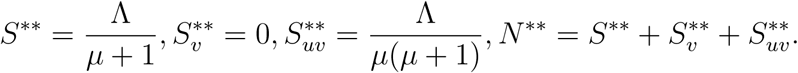

Clearly ℛ_*NPI*_ > ℛ_*A*_ which means that the non-pharmaceutical intervention will significantly reduce the community transmission and the rate of secondary infection. Similarly, in the absence of all non-pharmaceutical interventions, ie. setting all non-pharmaceutical intervention parameters *v*_1_ and *v*_3_ to zero in (17), we have

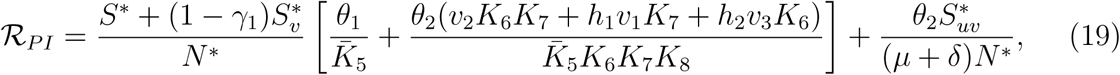

where 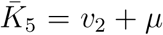. Clearly ℛ_*PI*_ > ℛ_*A*_ which also means that the pharmaceutical intervention will significantly reduce the community transmission and the rate of secondary infection. When strict social distancing measure is implemented, i.e. when *θ*_1_ = *θ*_2_ = 0 then ℛ_*A*_ = 0. This clearly means that strict social distancing can go a long way in reducing the generation of inter-community transmission of COVID-19 to zero. This is in confirmation with the recommendation of the World Health Organization [8, 20] and the work of [11].

To get the basic reproduction number without any control measures, we set all intervention parameters to zero i.e. *ρ* = *γ*_1_ = *f*_1_ = *f*_2_ = *v*_1_ = *v*_3_ = *S*_*v*_ = *S*_*uv*_ = 0, to have

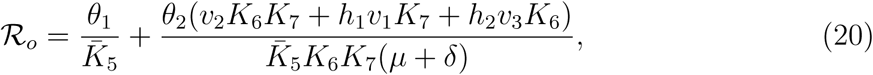

since *S*^∗^ = *N* ^∗^. It is easy to conclude that

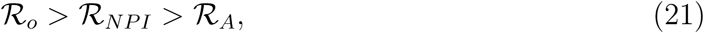

and

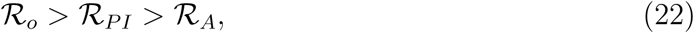

which is in confirmation and agreement with the work of [11]. Simulating all the results in this section using the parameter values and data presented under section 4 when *ρ* = *γ*_1_ = 0.8, the reproduction number in equation (17) when all the control measures are implemented gives 0.5521 (< 1 which is locally asymptotically stable). The one in equation (18) when only the non-pharmaceutical intervention is implemented gives 19.4827 (> 1 which is not locally asymptotically stable) while that of equation (19) when only the pharmaceutical intervention is implemented gives 0.8133 (< 1 which is locally asymptotically stable). When all interventions are set to zero as appeared in equation (20), the reproduction number becomes 19.6815 (> 1 which is not locally asymptotically stable). All these are in good agreement with the expressions in equation (21) and (22).

## 4 Numerical Simulation of The Impacts of Interventions and Model Fitting

Taking South Africa as a case study, we present the numerical simulation of the presented model equations (3)-(12) to understand the impacts of all intervention strategies.

### 4.1 Data Acquisition and Parameter Estimation

The detailed acquisition of the data and the estimation of model parameters using the officially published COVID-19 data within the period of July 1, 2020 and July 31, 2020 shall be extensively discussed here. The simple rationale behind the choice of taking South Africa as a case study is because South Africa has maintained the crown as the COVID-19 epicenter in Africa. The Figure 2 (reproduced in bar-chart form) shows the spread of the pandemic across several countries in the world.

**Fig. 1.**
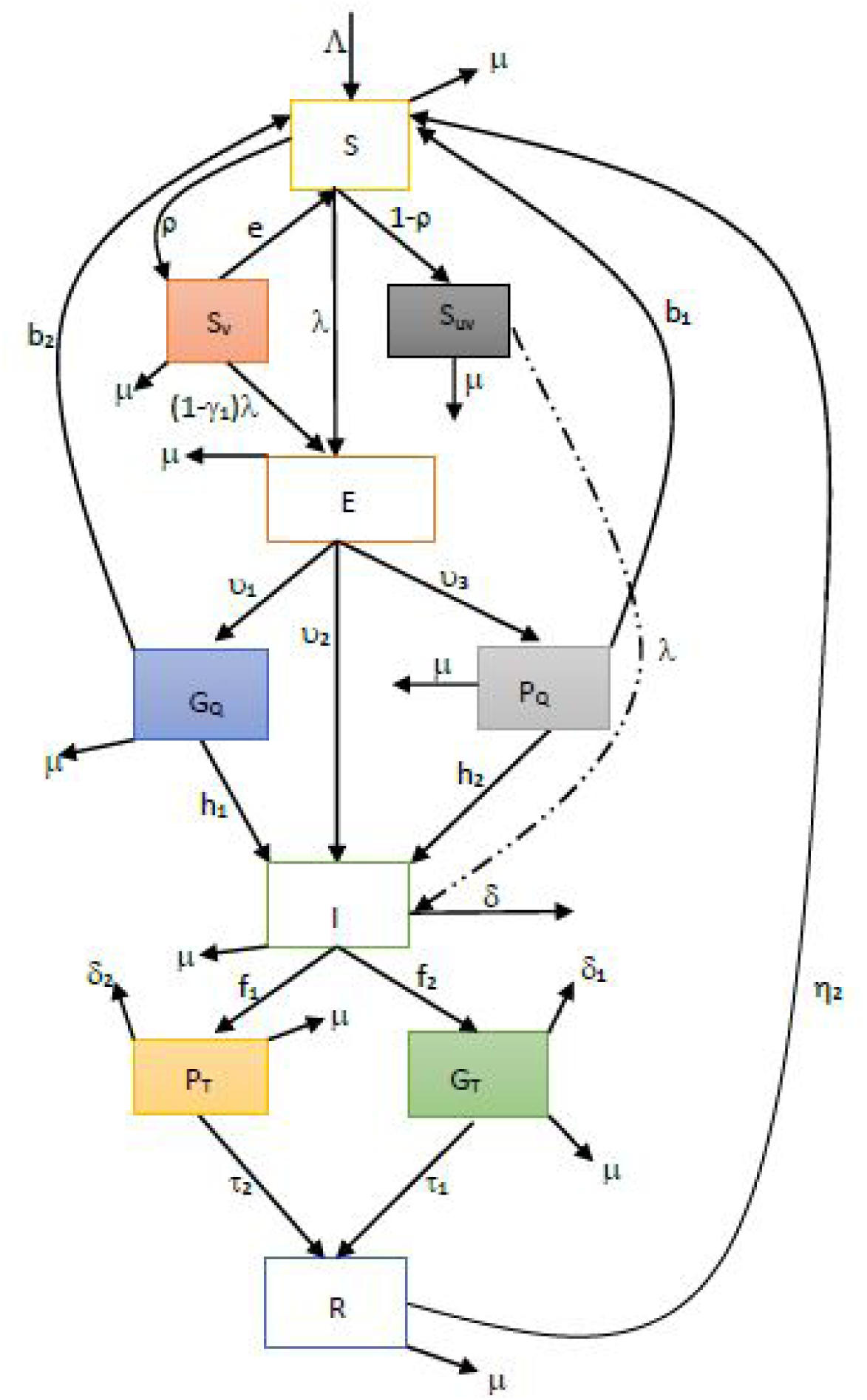
Flow chart of the model.

**Fig. 2.**
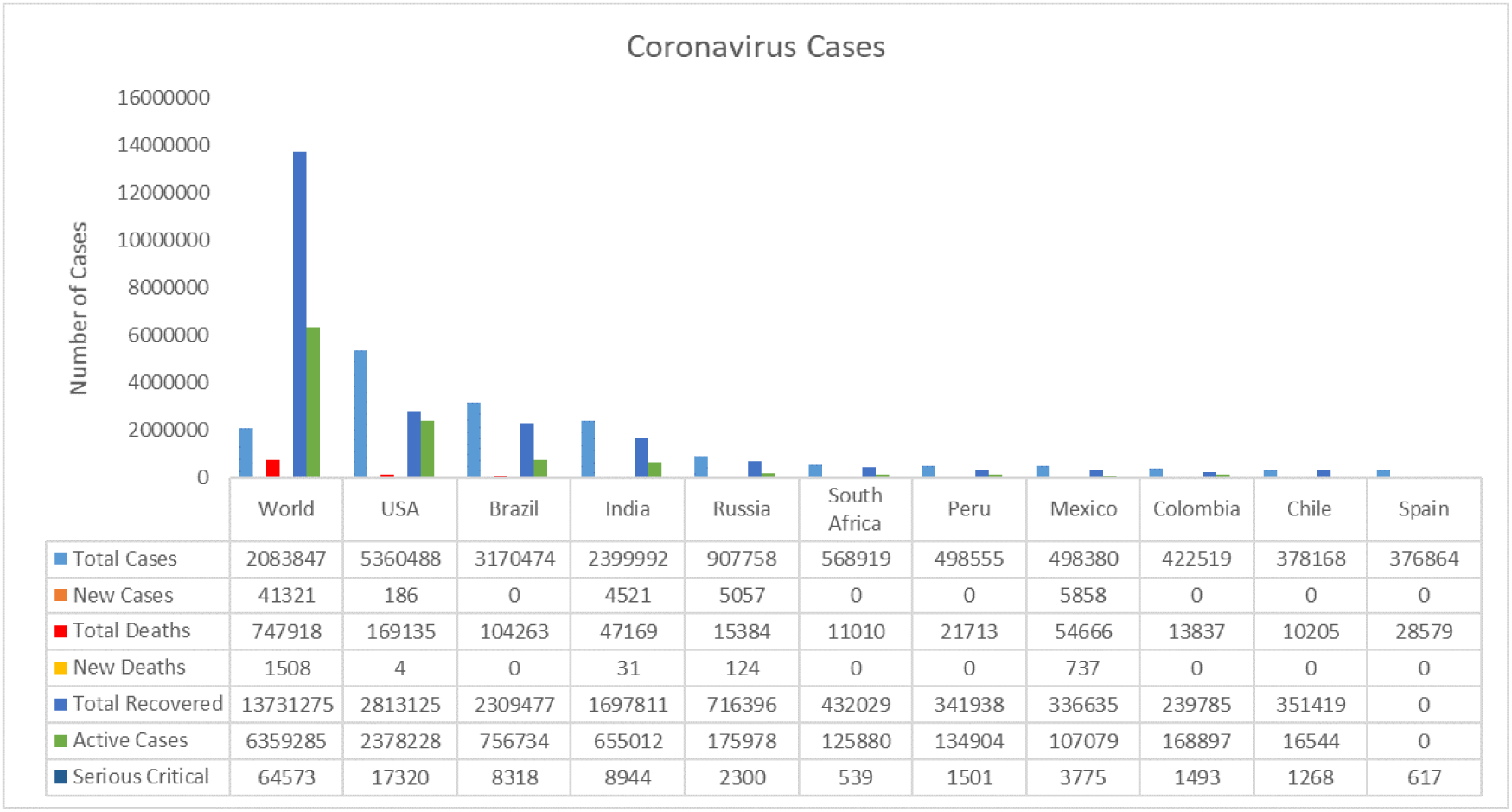
World COVID-19 Distribution [31].

As at July 31, 2020, the number of confirmed cases in South Africa was 493,183 [31] with 135,214 active cases which account for approximately 2.5% of the total world COVID-19 cases [31]. More worryingly, South Africa is currently ranked 5th among the most affected countries. In the month of July alone, South Africa recorded more than 60% of their total confirmed cases. The month of July, according to experts, is the peak of cases detection in the country and it really underlines the reason why the country is considered best choice for this research.

According to the South African mid-year population [25], the migration/recruitment rate is estimated at 18% of the total population which is 1,039,749 equivalent to 2,795 people per day. Hence, parameter Λ = 2, 795. It is also revealed from the data that South African life expectancy for both sexes is 64.88 years, therefore the natural death rate is given by 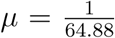. The total reported death from COVID-19 complications with the period of July 1, 2020 to July 31, 2020 is 5,256 which makes the death rate estimated at *δ* = 0.0157. Since the data for those that die under both personal and government treatments are not clearly differentiated, we assume that *δ*_1_ = 0.0112 and *δ*_2_ = 0.0142. It was also gathered that 71.% of infected people recovered so that the recovery rate is for people under treatment in a government owned hospital is 0.710 and we assume that *τ*_2_ = 0.0212 for those under personal treatment.

All other parameter values are extracted from available literature and referenced in Table 1. After fitting the South African data to the model, from table 2, we set *I* = 159, 333 for the number of detected and confirmed cases. Since no vaccine is available presently, we set *S*_*v*_ = *S*_*uv*_ = 0. All other state variables can not be determined from the available data as at the time of this research, so we chose the following

**Table 1:** Hypothetical Value of Parameters

| Parameter | Value (per day) | Source |
| --- | --- | --- |
| $\theta_1$ | 0.2189 | [18] |
| $\Lambda$ | 2795 | [25] |
| $\mu$ | $\frac{1}{64.88}$ | [31] |
| $f_1$ | 0.1824 | [18] |
| $f_2$ | 0.2337 | [18] |
| $\delta_1$ | 0.0112 | Estimated |
| $h_1$ | 0.0215 | Assumed |
| $h_2$ | 0.0255 | Assumed |
| $\delta_2$ | 0.0142 | Assumed |
| $\delta$ | 0.0157 | [31] |
| $\tau_1$ | 0.0710 | [31] |
| $\tau_2$ | 0.0212 | [31] |
| $\theta_2$ | 0.6123 | Estimated from [18] |
| $v_2$ | 0.1792 | [25] |
| $v_1$ | 0.1121 | Estimated from [18] |
| $\gamma_1, \rho$ | [0,1] | Variable |
| $b_1, b_2$ | 0.3521, 0.2151 | Assumed |
| $e, \eta_2, v_3$ | 0.0152, 0.2132, 0.4210 | Assumed |

**Table 2:** Cumulative Number of Reported Cases For The Month of July

| Day | Number of Cases | Day | Number of Cases |
| --- | --- | --- | --- |
| 1 | 159,333 | 16 | 324,221 |
| 2 | 168,061 | 17 | 337,594 |
| 3 | 177,124 | 18 | 350,879 |
| 4 | 187,977 | 19 | 360,550 |
| 5 | 196,750 | 20 | 373,628 |
| 6 | 205,721 | 21 | 381,798 |
| 7 | 215,855 | 22 | 394,948 |
| 8 | 224,665 | 23 | 408,052 |
| 9 | 238,339 | 24 | 421,996 |
| 10 | 250,687 | 25 | 434,200 |
| 11 | 264,184 | 26 | 445,433 |
| 12 | 276,242 | 27 | 452,529 |
| 13 | 287,796 | 28 | 459,761 |
| 14 | 298,292 | 29 | 471,123 |
| 15 | 311,049 | 30 | 482,169 |
|  |  | 31 | 493,183 |

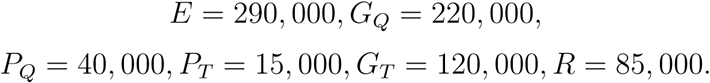

The South African population as at 2019 is estimated at 58,775,022 [25], therefore *N* = 58 × 10^6^ and hence

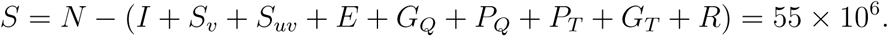

### 4.2 Model Fitting

To test the suitability of the model equations (3)-(12) to assess the impacts of both pharmaceutical and non-pharmaceutical interventions in curbing the global pandemic, we have extracted the monthly spread of the South African confirmed cases as appeared in figure 3. Since the month of July is the focus, we have consequently extracted the number of daily confirmed cases for the said month as appeared in Table 2.

**Fig. 3.**
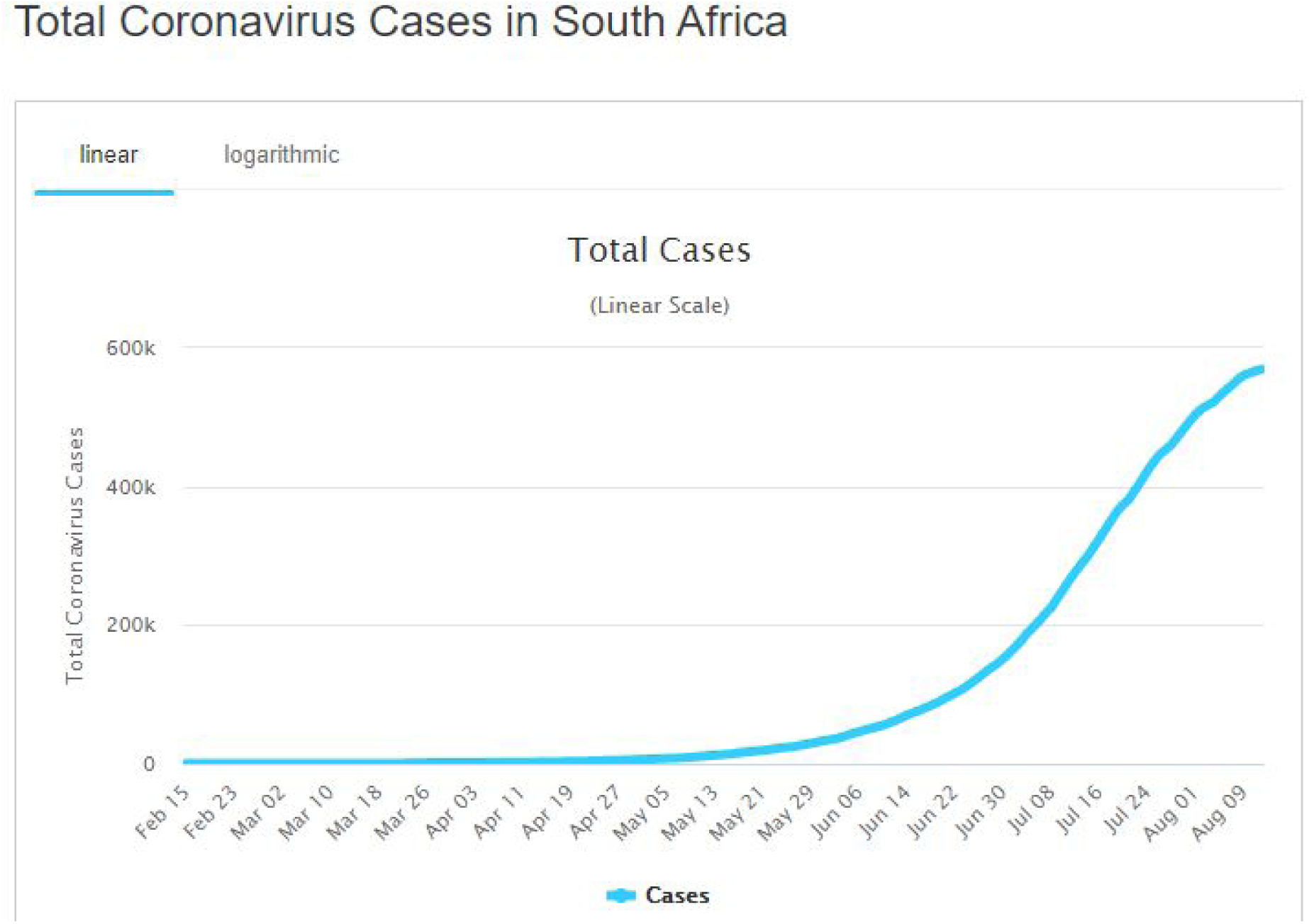
Monthly COVID-19 Distribution in South Africa [31] (Reproduced in Bar-Chart Form).

Using the earlier mentioned data, the equations (3)-(12) give a very good fit of the South African COVID-19 data for the period of July 1, 2020-July 31, 2020 as depicted in Figure 4. More-so, to examine the closeness of the model equations to the published data, Non-linear Least Squares (NLS) method shall be used [13].

**Figure 4:**
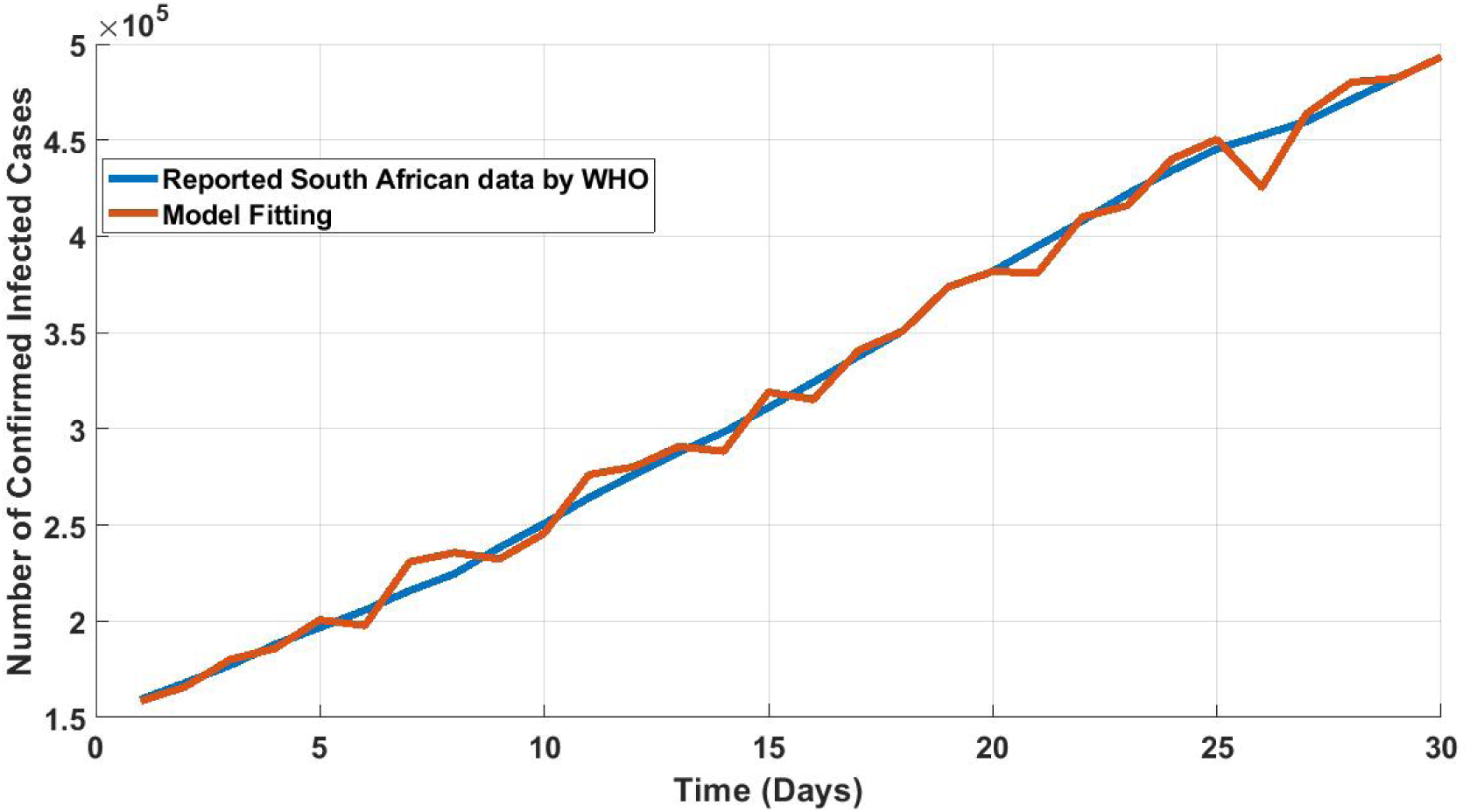
Comparison of observed South African COVID-19 data and model prediction for the period of July 1, 2020-July 31, 2020 with parameter values in Table 1 and 2.

Suppose *y*_*OD*_ denotes the published data. Then, the one predicted by the model given by *y*_*pred*_ is obtained using the equation:

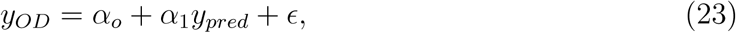

where *α*_*o*_ denotes the intercept and *α*_1_ denotes the slope of the regression line and *ϵ* represents the random error. If the coefficient of determination *R*^2^ = 1, *α*_0_ = 0 and *α*_1_ = 1, then the model is ideal. Using MATLAB, we obtained *α*_*o*_ = 0.0832 and *α*_1_ = 0.9707 (with their corresponding 95% confidence intervals [0.0360 0.1101] and [0.9281 0.9782], respectively) and *R*^2^ = 0.9991 using the aforementioned initial data and parameter values in Table 1 and 2. Thus, the OLS regression analysis established the closeness of the model and the real data. Hence, the model equations (3)-(12) are realistic and can be used to study the COVID-19 model and impacts of both interventions in South Africa.

### 4.3 Impacts of Social Distancing only on the Reproduction Number

The model (3)-(12) is firstly simulated for the special case where the only implemented intervention is social distancing. Doing this will make it convenient for us to examine the impact of social-distancing measures as the only anti-COVID-19 intervention strategy in the country. It is worth noting here that social-distancing is measured in terms of the reduction in the numerical values of *θ*_1_ and *θ*_2_ (known as contact rates) from their baseline values. The result obtained is presented in figure 5 in terms of the associated reproduction number.

**Figure 5:**
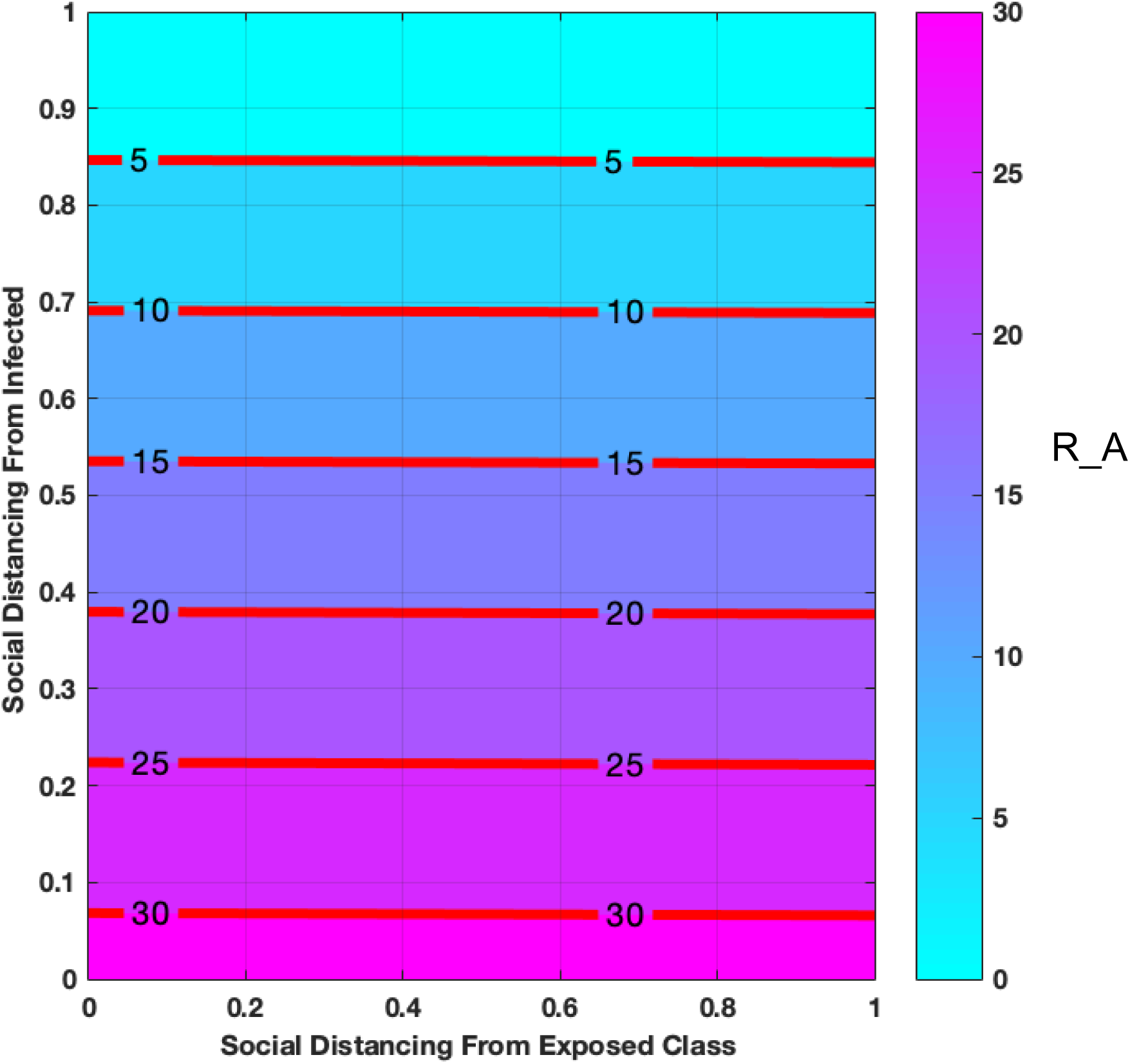
Impact of Social Distancing only on Reproduction Number.

We also have to mention here that the point where contact rates *θ*_1_ = *θ*_2_ = 0 corresponds to 100% level of social distancing while *θ*_1_ = *θ*_2_ = 1 corresponds to absolute abuse/absence of the social distancing intervention. This is because social distancing is a measure of reduction in contact rates. When contact rates reduce, social distancing is upheld and when contact rates is high, social distancing is abused. From the presented figure 5, when *θ*_1_ = *θ*_2_ = 0 (when social distancing is at 100%) the reproduction number which accounts for the rate at which community transmission occurs becomes zero. Also, when *θ*_1_ = *θ*_2_ = 1 (when social distancing is totally disregarded) the reproduction number becomes very high which is very dangerous in the fight against COVID-19. This is in confirmation with the recommendation of WHO regarding the importance of social distancing and lock-down.

### 4.4 Impacts of Social Distancing with Varying Level of Vaccination and Vaccine Efficacy on the Reproduction Number

In this subsection, we shall incorporate vaccination to the already established social distancing intervention. Figure 6 shows the effect of social-distancing with different level of vaccine and its efficacy.

**Figure 6:**
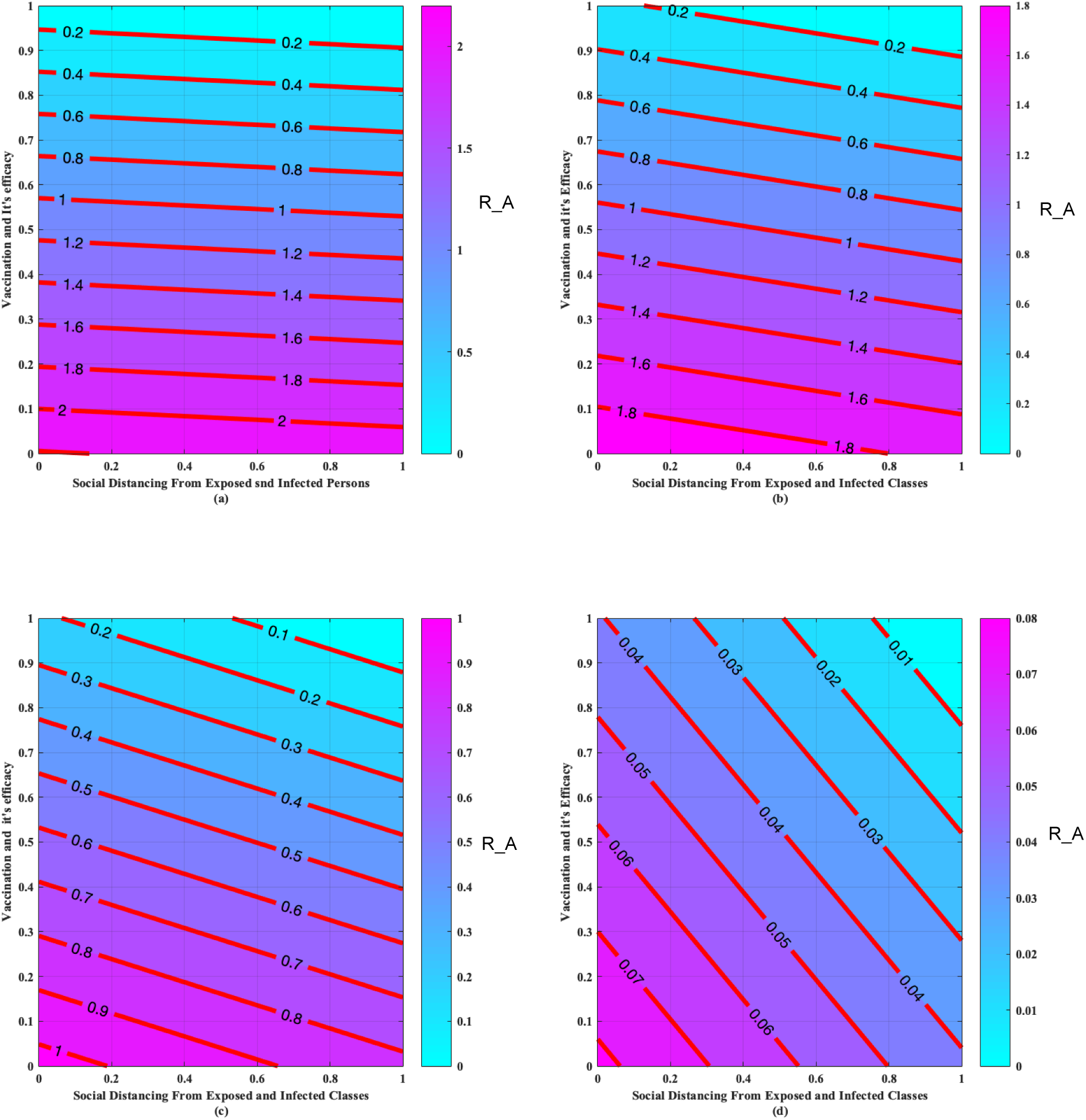
Effect of Social Distancing with (a) 10% level of vaccination and efficacy (b) 40% level of vaccination and efficacy (c) 80% level of vaccination and efficacy (d) 100% level of vaccination and efficacy.

It can be observed that the higher the social distancing with different level of vaccination and vaccine efficacy, the lower the COVID-19 transmission rate. In fact when the vaccination is at 0% the COVID-19 reproduction number was 2.2 in figure 6 (a) but when vaccination is at 100%, the community COVID-19 transmission reduced to 0.08 in figure 6(d) which should be enough to contain the spread of the dreadful pandemic. We also note in figure (5) that in the absence of social-distancing measure ie. when *θ*_1_ = *θ*_2_ = 1 the reproduction number was 30 but when vaccination was introduced at %100, the transmission rate dropped to 0.08 as depicted in figure 6 (d). This also underlines the effect of robust vaccination in the fight against the notorious pandemic.

### 4.5 Impacts of Vaccination with Varying Level of Anti COVID-19 Treatments on the Reproduction Number

We further incorporate both personal and government treatments to the already established vaccination programme. Figure 7 shows the effect of different level of treatments under vaccination intervention.

**Figure 7:**
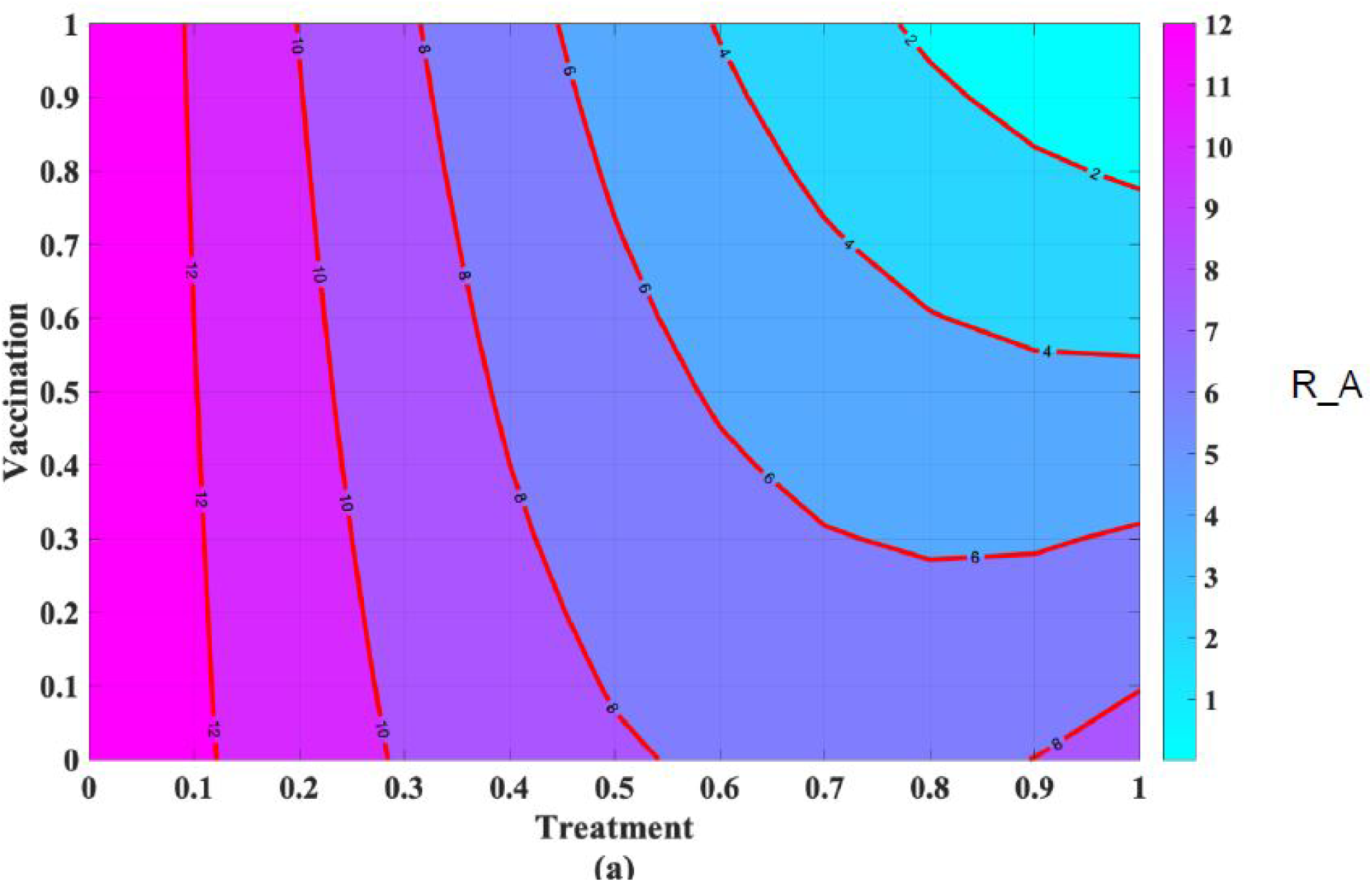
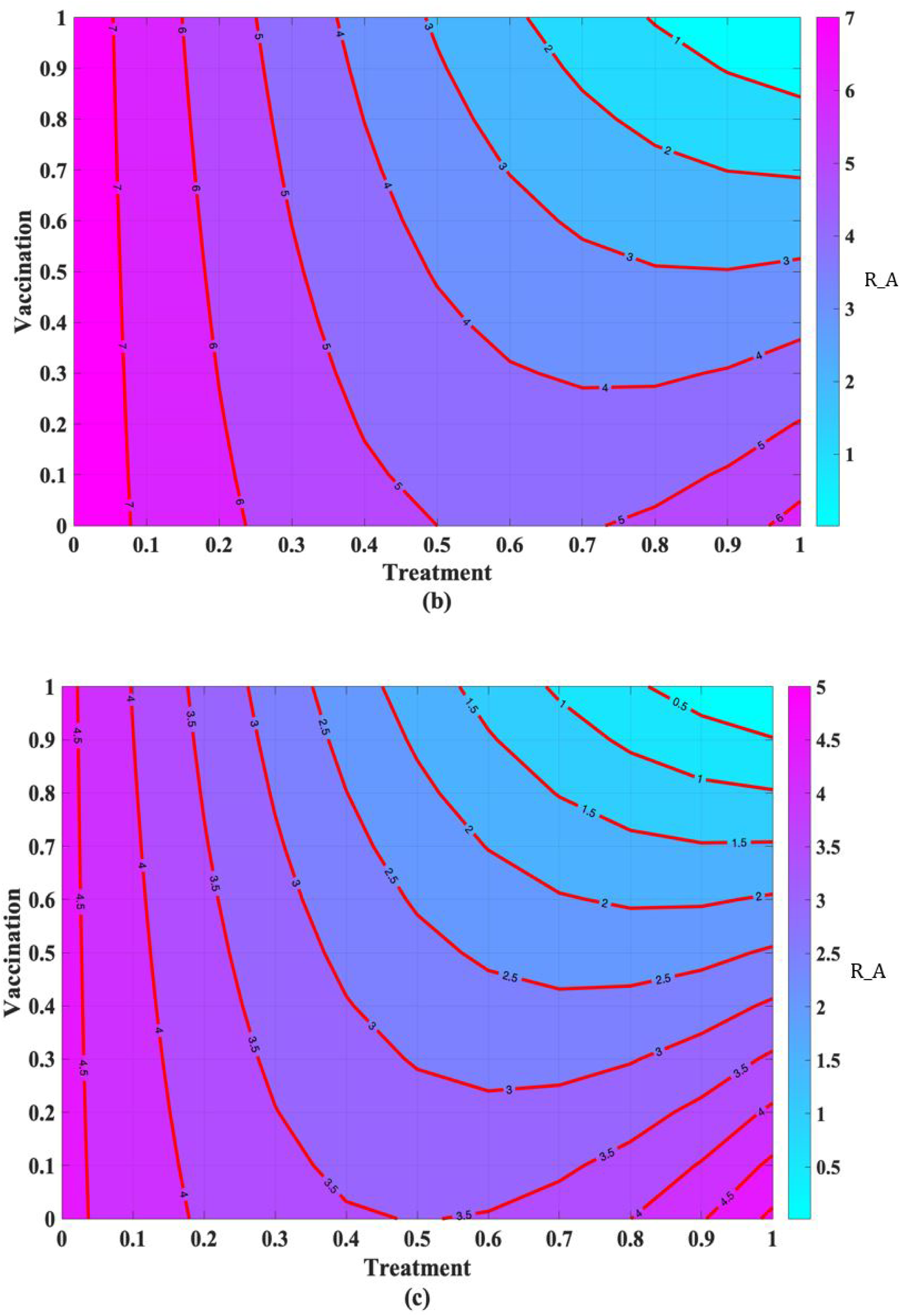
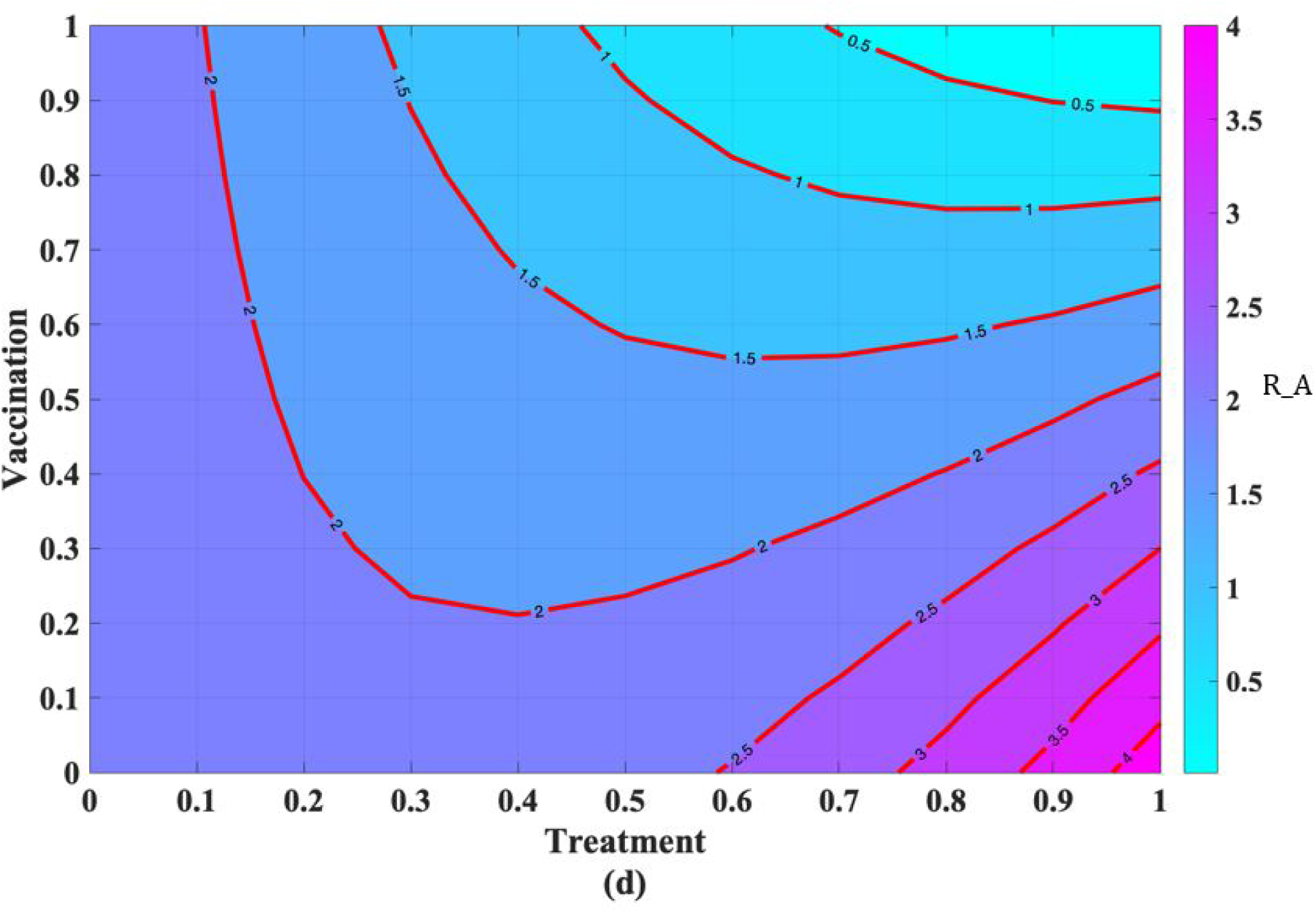
Effect of vaccination with (a) 10% level of treatment (b) 30% level of treatment (c) 50% level of treatment (d) 100% level of treatment.

It can be observed that the higher the vaccination and vaccine efficacy with different treatment levels, the lower the COVID-19 reproduction number. In fact when the treatment level is at 0% the COVID-19 reproduction number was 12 but when the treatment level is at 100%, the community COVID-19 transmission reduced to 4. This also underlines the importance of treatment and vaccination as recommended by the WHO.

### 4.6 Impacts of Both Interventions on the Reproduction Number

Finally we shall examine the combined effect of all the intervention strategies. We shall consider the combined effects of pharmaceutical interventions first followed by non-pharmaceutical interventions and then consider when both are concurrently implemented to combat the pandemic.

Figure 8 clearly shows the importance of both intervention in curbing the pandemic. The figures suggest that as much as the non-pharmaceutical interventions such as social distancing, lock-down and face mask are important, the impacts of the pharmaceutical interventions such as treatment and vaccination is absolutely incomparable in curbing the disease. Figure 8 (c) vividly shows that both intervention will reduce the community transmission to almost zero within a very short period of time.

**Figure 8:**
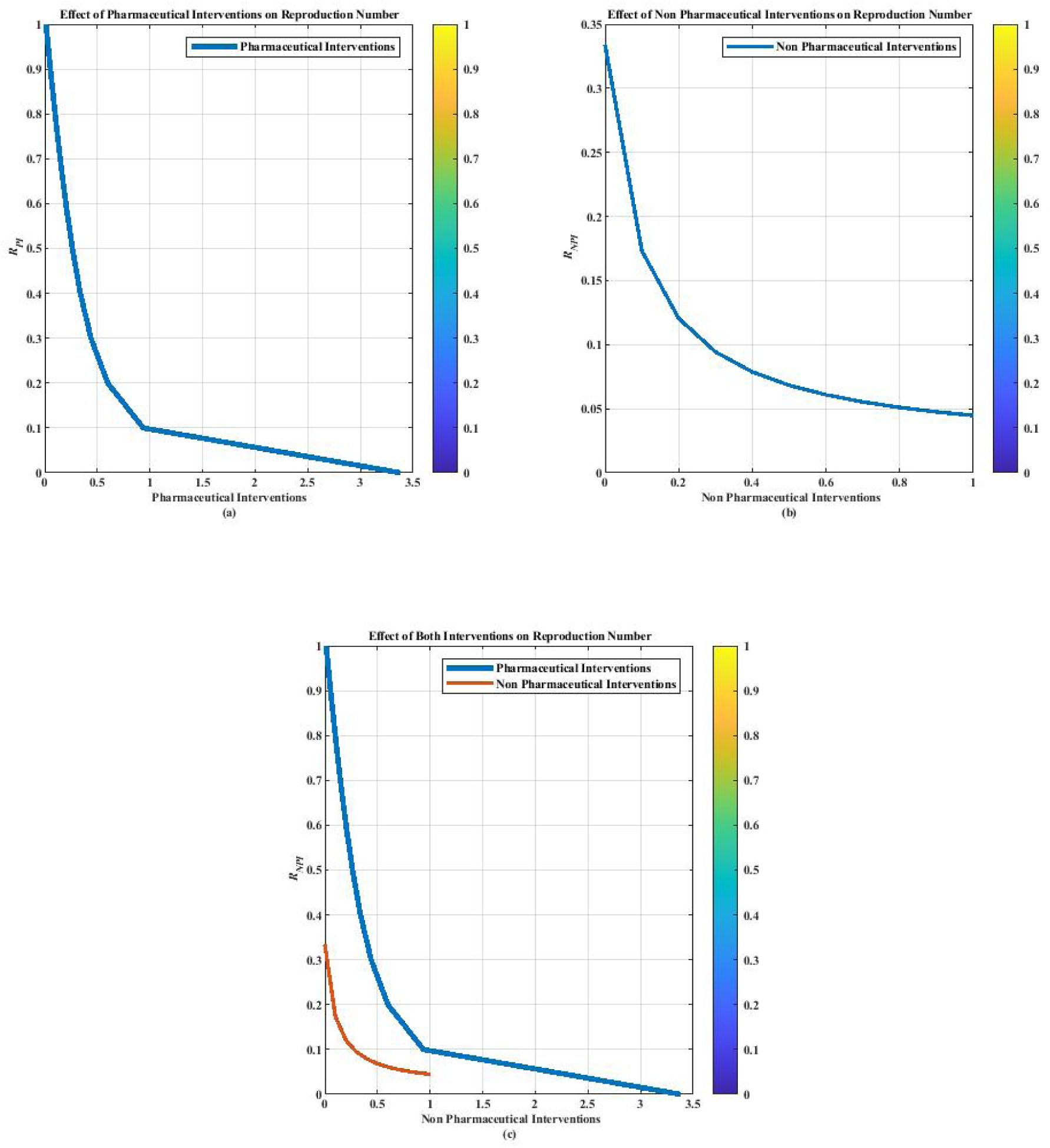
Impacts of Both Interventions on the Reproduction Number.

## 5 Results and Discussion

A novel COVID-19 was first detected in Wuhan, China in December 2019 which has infected 21,388,904 persons with more than 764,17 deaths across the globe as at August 15, 2020 at 3:00 pm [31]. Neither effective treatment nor vaccine is available at the moment, attentions are consequently shifted to non pharmaceutical interventions such as maintenance of physical/social-distancing, use of nose-mask, face-shield, lock-down enforcement, restriction of movement of non essential workers, shut down of economy, quarantine of exposed persons, isolation of infected individuals among others. Efforts are also in advanced stage that an anti-COVID 19 vaccine will be globally approved sooner rather than later.

After establishing the basic properties of the model which include positivity solution and invariant region, we established the condition for local stability of the virus-free equilibrium (VFE) and we claimed that the VFE is stable when the associated threshold parameters is less than unity. The Theorem was stated which can be proved using Theorem 2 of [26]. The biological implication of this is that the novel corona-virus will die out of the community whenever the associated threshold parameters ℛ_*o*_ < 1, ℛ_*NPI*_ < 1, ℛ_*PI*_ < 1 as the case maybe. In other words, if both recommended interventions (ie. social distancing, quarantine, treatment and vaccination) are fully implemented, the threshold parameters can be brought down to less than unity and consequently ensure the eradication of the pandemic. This has been simulated under equations (19)-(22) as mentioned earlier.

Using the officially published South African COVID-19 data, the model equations (3)-(12) gave a very good fit of the South African COVID-19 data for the period of July 1, 2020-July 31, 2020 as depicted in Figure 4. Furthermore, to qualitatively assess the closeness of the model against the real data, we employed the Non-linear Least Squares (NLS) method. We used MATLAB to obtain *α*_*o*_ = 0.0832 and *α*_1_ = 0.9707 (with their corresponding 95% confidence intervals [0.0360 0.1101] and [0.9281 0.9782], respectively) and *R*^2^ = 0.9991 using the aforementioned initial data and parameter values in Table 1 and 2.

Several reproduction numbers were calculated such as the basic reproduction number ℛ_*o*_, the reproduction number for pharmaceutical intervention only ℛ_*PI*_ and the reproduction number for non-pharmaceutical intervention only ℛ_*NPI*_. We also explicitly explain that the reproduction number (which is the measure of average number of new virus infection developed by a single COVID-19 infected individual in a community) is low (which makes the virus transmission controllable) when intervention programmes are adequately implemented and high when no intervention is implemented. This kind of analysis is missing in majority of the published COVID-19 models. Though it was partly analyzed in [11] which was not as detailed as the one presented here and obviously missing in [15],[17],[18] and [20].

The model was first simulated to obtain the effects of social distancing as the only intervention strategy, while the other intervention strategies are set to zero. The result as depicted in figure 5 shows that the reproduction number drastically reduced from 30 to 5 as the physical distancing is adequately implemented in the country. The reproduction number consequently becomes zero when the physical distancing is maximum i.e (when *θ*_1_ = *θ*_2_ = 0) and becomes 30 when no physical distancing is upheld and free mixing, social gathering, school activities, social lives are allowed to continue as normal. This in confirmation with the recommendation of the World Health Organization (WHO) [27, 28, 29, 30], and the work of [11].

We further examined the impacts of social distancing with different vaccination level and vaccine efficacy on the Reproduction Number in order to grasp the level of vaccination and vaccine efficacy needed to curb the dynamics in South Africa. We considered the effects of Social Distancing with 10%, 40%, 80% and 100% levels of vaccination and efficacy. It can be seen that when social distancing remains constant, the reproduction rate of new infected confirmed cases reduces drastically as the vaccine coverage and efficacy increases. According to figures 6 (a,b,c,d), when the vaccination is at 0% the COVID-19 reproduction number was 2.2 in figure 6 (a) but when vaccination is at 100%, the community COVID-19 transmission reduced to 0.08 in figure 6(d). Though it seems extremely difficult to obtain this level of vaccine coverage and efficacy for many reasons. This is because sufficient vaccine will be less available at the early stage of the vaccination process which may not be enough to cover a country of almost 60 million population like South Africa. The vaccine, at early production stage, may still be prone to some clinical errors needed to be rectified as time goes on. So vaccination level and efficacy of upto 80%, 90% etc for almost 60 million population will seriously take a long period of time before it can be achievable. That is the more reason why the simulation complemented vaccination coverage with treatment and other non-intervention strategies like social distancing.

The impacts of vaccination with different treatment levels of anti-COVID-19 treatment on the Reproduction Number was also examined. It can be observed that the higher the vaccination and vaccine efficacy with different treatment levels, the lower the COVID-19 reproduction number. In fact, when the treatment level is at 0% the COVID-19 reproduction number was 12, but when the treatment level is at 100%, the community COVID-19 transmission reduced to 4 which accounts for almost 70% reduction in community transmission. This also underlines the importance of treatment and vaccination as recommended by the WHO.

## 6 Concluding Remarks

According to the World Health Organization (WHO), the causative agent of the novel COVID-19 is Severe Acute Respiratory Syndrome Corona-virus 2 (SARS-CoV-2) that was first discovered in Wuhan, China in December 2019. Within a very short period of time, the virus has claimed lives of thousands of infected persons while more than 2 million persons are hospitalized.

While researchers are still busy in the laboratories to develop vaccine to curb the menace, attentions are shifted to non-pharmaceutical controls to mitigate the burden of the novel pandemic. To tackle this challenge, a ten compartmental mathematical model was developed and presented in this paper to investigate the possible effects of both pharmaceutical and non-pharmaceutical controls such as self quarantine and treatments as well as government’s provided quarantine and treatment facilities. Several reproduction numbers were calculated and used to determine the impact of both control measures as well as projected benefits of social distancing, treatments and vaccination on the community transmission.

We considered the impacts of social distancing only on the Reproduction Number. We also investigated and compared the possible impact of social distancing with different levels of vaccination on the Reproduction Number. Furthermore, we examined the impacts of vaccination while different levels of treatment was administered. Using the officially published South African COVID-19 data, the numerical simulation showed that the community reproduction number will be 30 when there is no social distancing but will drastically reduced to 5 when social distancing is adequately implemented to some certain degree of compliance. This accounts for more than 80% reduction in disease transmission. More so, when vaccination with good efficacy was implemented, the community reproduction number was 4 which increases to 12 without vaccination. This accounts for almost 70% reduction in disease transmission.

Finally, we established that the implementation of both interventions is enough to curtail the spread of the novel coronavirus disease in South Africa, which is in confirmation with the recommendation of the world health organization and provided a better insight to the pandemic than several other existing research work such as [11] (whose model is less detailed) and [12] (whose work does not involve both interventions we considered).

## Data Availability

All data used is available within the body of the manuscript

## References

[1] Bai, Y., Yao, L., Wei, T., Tian, F., Jin, D. Y., Chen, L., & Wang, M. (2020). Presumed asymptomatic carrier transmission of COVID-19. Jama, 323(14), 1406–1407.

[2] Bryner J., Ghose T., Rettner R., Saplakoglu Y., Lanese N., Coronavirus cases top 94,000: Live updates on COVID-19, Live Sci. (2020) (Accessed on 4 August 2020).

[3] Callaway E., “Scores of coronavirus vaccines are in competition — how will scientists choose the best?” a natureresearch journal (Assessed on April 30, 2020).

[4] Centers for Disease Control, Prevention, Coronavirus Disease 2019 (COVID-19), National Center for Immunization and Respiratory Diseases (NCIRD), Division of Viral Diseases, 2020, https://www.cdc.gov/coronavirus/2019-ncov/index.html (Accessed on 4 August 2020).

[5] Del Rio, C., & Malani, P. N. (2020). COVID-19—new insights on a rapidly changing epidemic. Jama, 323(14), 1339–1340.

[6] Diekmann, O., Heesterbeek, J. A. P.,& Metz, J. A. (1990). On the definition and the computation of the basic reproduction ratio ℛ_0_ in models for infectious diseases in heterogeneous populations. Journal of mathematical biology, 28(4), 365–382.

[7] Dong, E., Du, H., & Gardner, L. (2020). An interactive web-based dashboard to track COVID-19 in real time. The Lancet infectious diseases, 20(5), 533–534.

[8] Ferguson, N., Laydon, D., Nedjati Gilani, G., Imai, N., Ainslie, K., Baguelin, M., … & Dighe, A. (2020). Report 9: Impact of non-pharmaceutical interventions (NPIs) to reduce COVID19 mortality and healthcare demand.

[9] Garba, S. M., Safi, M. A., & Usaini, S. (2017). Mathematical model for assessing the impact of vaccination and treatment on measles transmission dynamics. Mathematical Methods in the Applied Sciences, 40(18), 6371–6388.

[10] Garba, S.M., Lubuma J.M.S. and Tsanou B., Modeling the transmission dynamics of the COVID-19 Pandemic in South Africa, Mathematical Biosciences (2020), doi: https://doi.org/10.1016/j.mbs.2020.108441

[11] Gumel, A. B., Iboi, E. A., & Ngonghala, C. N. (2020). Will an imperfect vaccine curtail the COVID-19 pandemic in the US?. medRxiv.

[12] Jia, J., Ding, J., Liu, S., Liao, G., Li, J., Duan, B.,& Zhang, R. (2020). Modeling the Control of COVID-19: Impact of Policy Interventions and Meteorological Factors. Electronic Journal of Differential Equations, 2020(23), 1–24.

[13] Kendall M.G. and Stuart A. The Advanced Theory of Statistics, volume 2. Inference and Relationship London:Charles Gri±n, 1979.

[14] Kirkpatrick D., “In race for a Coronavirus vaccine, an Oxford Group Leaps Ahead,” New York Times (May 2, 2020) https://www.nytimes.com/2020/04/27/world/europe/coronavirus-vaccine-update-oxford.html.

[15] Kucharski AJ, Russell TW, Diamond C, et al. Early dynamics of transmission and control of COVID-19: a mathematical modelling study. Lancet Infect Dis 2020. https://doi.org/10.1016/S1473-3099(20)30144-4.

[16] Lin Q, Zhao S, Gao D, et al. A conceptual model for the outbreak of Coronavirus disease 2019 (COVID-19) in Wuhan, China with individual reaction and governmental action. Int J Infect Dis 2020;93:211–6. https://doi.org/10.1016/j.ijid.2020.02.058.

[17] Li, Q., Guan, X., Wu, P., Wang, X., Zhou, L., Tong, Y.… & Xing, X. (2020). Early transmission dynamics in Wuhan, China, of novel coronavirus–infected pneumonia. New England Journal of Medicine.

[18] Mushayabasa, S., Ngarakana-Gwasira, E. T., & Mushanyu, J. (2020). On the role of governmental action and individual reaction on COVID-19 dynamics in South Africa: A mathematical modelling study. Informatics in Medicine Unlocked, 100387.

[19] National Institute for Communicable Diseases. First Case of COVID-19 Coronavirus Reported in South Africa. https://www.nicd.ac.za/first-case-of-covid-19-coronavirus-reported-in-sa/.

[20] Ngonghala, C. N., Iboi, E., Eikenberry, S., Scotch, M., MacIntyre, C. R., Bonds, M. H., & Gumel, A. B. (2020). Mathematical assessment of the impact of non-pharmaceutical interventions on curtailing the 2019 novel Coronavirus. Mathematical Biosciences, 108364.

[21] Rabiu, M., Willie, R., & Parumasur, N. (2020). Mathematical analysis of a disease-resistant model with imperfect vaccine, quarantine and treatment. Ricerche di Matematica, 1–25.

[22] Rabiu Musa, Robert Willie, Nabendra Parumasur, Analysis of a virus-resistant HIV-1 model with behavior change in non-progressors, Biomath 9 (2020), 2006143, http://dx.doi.org/10.11145/j.biomath.2020.06.143

[23] Silverstein W.K., Stroud L., Cleghorn G.E., Leis J.A., First imported case of 2019 novel coronavirus in Canada, presenting as mild pneumonia, Lancet 395 (10225) (2020) 734.

[24] South Africa Begins Nationwide Coronavirus Lockdown.www.voanews.com/science-health/coronavirus-outbreak/south-africa-begins-nationwide-coronavirus-lockdown#:~:text=South%20African%20Defense%20Forces%20patrol,coronavirus%20lockdown%20March%2027%2C%202020.&text=South%20Africans%20on%20Friday%20began,topping%20900%20in%20South%20Africa.

[25] The South African mid-year population (2019). Available on:http://www.statssa.gov.za/publications/P0302/P03022019.pdf.

[26] Van den Driessche, P., & Watmough, J. (2002). Reproduction numbers and sub-threshold endemic equilibria for compartmental models of disease transmission. Mathematical biosciences, 180(1-2), 29–48.

[27] World Health Organization, Coronavirus disease (COVID-19) technical guidance, WHO (2020) https://www.who.int/emergencies/diseases/novel-coronavirus-2019/technical-guidance (Accessed on 4 August 2020).

[28] World Health Organization WHO releases guidelines to help countries maintain essential health services during the COVID-19 pandemic. https://www.who.int/news-room/detail/30-03-2020-who-releases-guidelines-to-help-countries-maintain-essential-health-services-during-the-covid-19-pandemic.

[29] World Health Organization, Emergencies, preparedness, response. Pneumonia of unknown origin – China, Dis. Outbreak News (2020) https://www.who.int/csr/don/05-january-2020-pneumonia-of-unkown-cause-china/en/. (Accessed on 5 August 2020).

[30] World Health Organization. (2020). Coronavirus disease 2019 (COVID-19): situation report, 88.

[31] Worldometers. Coronavirus cases. Online, https://www.worldometers.info/coronavirus/#countries; 2020 (Accessed 08.13.20.).

[32] Xu, X. W., Wu, X. X., Jiang, X. G., Xu, K. J., Ying, L. J., Ma, C. L., … & Sheng, J. F. (2020). Clinical findings in a group of patients infected with the 2019 novel coronavirus (SARS-Cov-2) outside of Wuhan, China: retrospective case series. bmj, 368.

[33] Yin, Y., & Wunderink, R. G. (2018). MERS, SARS and other coronaviruses as causes of pneumonia. Respirology, 23(2), 130–137.

[34] Zhou T, Liu Q, Yang Z, et al. Preliminary prediction of the basic reproduction number of the Wuhan novel coronavirus 2019-nCoV. Evid Base Med 2020;13(1):3–7.https://doi.org/10.1111/jebm.12376

